# Psychosis in Alzheimer’s Disease is Associated with Excitatory Neuron Vulnerability and Post-Transcriptional Mechanisms Altering Synaptic Protein Levels

**DOI:** 10.1101/2021.09.07.21262904

**Authors:** MR DeChellis-Marks, Y Wei, Y Ding, CM Wolfe, JM Krivinko, ML MacDonald, OL Lopez, RA Sweet, J Kofler

## Abstract

Alzheimer’s disease with psychosis (AD+P) is a phenotypic variant of the disease which is associated with a much more rapid deterioration compared to Alzheimer’s disease without psychosis (AD-P). The neurobiological basis of AD+P is poorly understood. AD is thought to be a disease of the synapse, and our previous studies suggest that those with AD+P have a differentially affected synaptic proteome relative to those with AD-P. We previously demonstrated that multiple neuropathologies only account for approximately 18% of the variance in the occurrence of psychosis in AD. In this study, we utilized RNA-sequencing of dorsolateral prefrontal cortex (DLPFC) in a cohort of 80 AD cases to evaluate novel transcriptomic signatures that may confer risk of psychosis in AD. We found that AD+P was associated with a 9% reduction in excitatory neuron proportion compared to AD-P [Mean (SD) AD+P 0.295 (0.061); AD-P 0.324 (0.052), p = 0.026]. Network analysis identified altered expression of gene modules from protein ubiquitination, unfolded protein response, eukaryotic initiation factor 2 (EIF2) signaling and endoplasmic reticulum stress pathways in AD+P. Including cell type proportions and differentially expressed modules with neuropathology measures explained 67.5% of the variance in psychosis occurrence in our AD cohort.

## Introduction

Psychosis in Alzheimer’s disease (AD with psychosis, AD+P), defined as the presence of delusions and/or hallucinations, is a distinct phenotype, comprising 40% - 60% of those affected by AD (Sweet et al., 2003). AD+P is associated with hastened cognitive decline and elevated mortality compared to AD without psychosis (AD-P) (Murray et al., 2014). AD+P patients endure greater cognitive impairment, are more likely to be institutionalized during illness, and are affected by neuropsychiatric disturbances (e.g. aggression, agitation, depression) at higher rates compared to AD-P counterparts (Gilley et al., 1991; Sweet et al., 2001). Because psychosis in AD is heritable, it is likely to have a distinct neurobiology (Bacanu et al., 2005; Barral et al., 2015; DeMichele-Sweet et al., 2018; Sweet et al., 2010; Sweet et al., 2002). Further elucidating the neurobiological basis of risk towards psychosis in AD may therefore provide leads to innovative preventions or treatments.

AD neuropathology is defined by fibrillar deposits of amyloid beta and phosphorylated tau. In most cases, however, one or more comorbid neuropathologies (Kapasi et al., 2017), including Lewy bodies (Hamilton, 2000), Transactive response DNA Binding Protein 43 kDa (TDP-43) inclusions (Tremblay et al., 2011), and cerebrovascular lesions are also present (Sarro et al., 2017; Snyder et al., 2015). Previously we set out to establish a comprehensive model of the association of these pathologies with psychosis risk in AD. This stepwise logistical regression identified pathologies that significantly associated with psychosis, including: phosphotau burden, presence of TDP-43 inclusions, an index of microglial activation, and indices of ischemia. However, these neuropathologies only accounted for 18% of the variance in psychosis risk (Krivinko et al., 2018). Further, we reported that after accounting for the contribution of neuropathologies, AD+P was associated with lower levels of canonical postsynaptic density proteins in the DLPFC compared to AD-P (Krivinko et al., 2018). Synaptic protein levels are potentially impacted by multiple factors including expression levels of their corresponding mRNAs and rates of translation initiation and of protein degradation. In the context of neurodegenerative disease, synaptic protein levels are also strongly impacted by neuron loss (Sweet et al., 2016).

Reduced gray matter volume and reduced indices of synaptic function in AD+P relative to AD-P has been replicated across multiple cerebral neocortical regions, particularly in bilateral frontal and prefrontal cortices (Murray et al., 2014). In this study we focused on the dorsolateral prefrontal cortex (DLPFC) of AD+P and AD-P subjects. We utilized RNA-sequencing to identify molecular mechanisms underlying the excess neuropathologic burden and altered synaptic proteostasis in AD+P. We found that excitatory neurons, as measured by unique excitatory neuron transcript levels, are selectively vulnerable in AD+P compared to AD-P. In addition, we identified co-expressed mRNA modules that were differentially expressed between AD+P and AD-P when controlling for cell proportion and were enriched for mRNAs regulating protein availability. Our findings suggest that the prior observation of synaptic protein reduction in AD+P relative to AD-P subjects results from contributions of neuronal survival and post-transcriptional regulation.

## Methods

### Subjects

We studied a cohort of 80 Alzheimer’s disease subjects (Table 1; Supplemental Methods: Appendix 1) obtained through the brain bank of the Alzheimer’s Disease Research Center (ADRC) at the University of Pittsburgh. All subjects, when living provided informed consent to participate using protocols approved by the University of Pittsburgh Institutional Review Board. The study of postmortem tissue was approved by the University of Pittsburgh Committee for Oversight of Research and Clinical Training Involving Decedents. Subjects underwent comprehensive evaluations by experienced clinicians in the University of Pittsburgh ADR), including neurologic, neuropsychological, and psychiatric assessments as previously described (Lopez et al., 2013; Sweet et al., 2000). Psychosis was evaluated with the CERAD behavioral rating scale (CBRS) (Tariot PN, 1995). The CBRS was administered at initial and annual visits and in some subjects between annual visits by telephone (DeMichele-Sweet et al., 2011; Sweet et al., 2010). Subjects were classified as AD+P if they had any hallucination or delusion symptom (CBRS item # 33-45) for 3 or more days in the previous month at any visit (DeMichele-Sweet et al., 2011; DeMichele-Sweet et al., 2018; Weamer et al., 2016; Weamer et al., 2009; Wilkosz et al., 2007; Wilkosz et al., 2006). Subjects with a preexisting psychotic disorder (e.g. schizophrenia) were excluded from the study. Statistical analyses of demographic and clinical differences between groups used t-test or Chi-Squared test as appropriate.

**Table 1.** Demographic, Clinical, and Tissue Characteristics of Subjects with Alzheimer’s Disease with and without Psychosis Examined by RNA-Sequencing.

| Variable | Alzheimer's Disease Without Psychosis<br>(n = 33) |  | Alzheimer's Disease With Psychosis<br>(n = 47) |  |
| --- | --- | --- | --- | --- |
|  | Mean or Total | SD or % | Mean or Total | SD or % |
| Age (Years) | 83.7 | $\pm 7.3$ | 82.0 | $\pm 6.0$ |
| Age at Onset (Years)<sup>a</sup> | 75.6 | $\pm 8.0$ | 72.1 | $\pm 6.6$ |
| Duration of Illness (Years)<sup>a</sup> | 8.1 | $\pm 3.0$ | 9.0 | $\pm 3.3$ |
| Postmortem Interval (hours) | 6.3 | $\pm 4.0$ | 5.9 | $\pm 3.9$ |
| Tau Area Ratio | 0.1 | $\pm 0.1$ | 0.1 | $\pm 0.1$ |
| Microvascular Lesion Sum Score | 0.3 | $\pm 0.5$ | 0.2 | $\pm 0.4$ |
| HLA-DR:Iba1 Ratio | 0.5 | $\pm 0.7$ | 0.4 | $\pm 1.3$ |
| Sex |  |  |  |  |
| Male | 21 | 63.6% | 28 | 59.6% |
| Female | 12 | 36.4% | 19 | 40.4% |
| Braak Stage |  |  |  |  |
| III | 5 | 15.2% | 5 | 10.6% |
| IV | 15 | 45.5% | 18 | 38.3% |
| V | 13 | 39.3% | 24 | 51.1% |
| APOE-4 Status |  |  |  |  |
| Positive | 17 | 51.5% | 29 | 61.7% |
| Negative | 16 | 48.5% | 18 | 38.3% |
| TDP-43 Pathology |  |  |  |  |
| Positive | 18 | 56.2% | 34 | 72.3% |
| Negative | 15 | 43.8% | 13 | 27.7% |
| Lewy Body Pathology<sup>b</sup> |  |  |  |  |
| Positive | 12 | 36.4% | 30 | 63.8% |
| Negative | 21 | 63.6% | 17 | 36.2% |
| Microvascular Lesion Count |  |  |  |  |
| 0 | 23 | 67.7% | 36 | 76.7% |
| $\geq 1$ | 10 | 32.3% | 11 | 23.4% |
| Antipsychotic Use |  |  |  |  |
| Yes | 2 | 6.1% | 9 | 19.1% |
| No | 31 | 93.9% | 38 | 80.9% |

<sup>a</sup>Data unavailable for one subject, Alzheimer's Disease Without Psychosis
<sup>b</sup>p < 0.05

### Sample Collection and Neuropathological Assessment

For ADRC subjects, frozen gray matter samples from the right superior frontal gyrus of the DLPFC were retrieved from the ADRC neuropathology core RNA sequencing. The corresponding formalin-fixed left DLPFC was used for immunostaining and processed for neuropathologic studies as previously described ((Krivinko et al., 2018); Supplemental Methods: Appendix 2). Neuropathologic diagnoses of Alzheimer disease were made according to NIA-Reagan criteria (Hyman and Trojanowski, 1997), with all cases meeting criteria for intermediate to high probability that their dementia was due to AD.

### Quantitative Immunohistochemistry and Digital Image Analysis

Neuropathological disease burden in the DLPFC was previously assessed in all 80 cases using quantitative immunohistochemistry (Krivinko et al., 2018). In short, serial 5 μm thick formalin-fixed, paraffin-embedded tissue sections were immunostained on an automated stainer (Discovery Ultra, Ventana, Tucson, AZ) using the following primary antibodies (Supplemental Methods Appendix 3): PHF-1, oligomeric tau T22, beta-amyloid NAB228, microglial markers Iba1 (Ionized calcium binding adaptor molecule 1), and HLA-DR (Human Leukocyte Antigen – DR isotype). No counterstaining was performed to ease signal quantification.

Whole slide digital images of the immunostained sections were created using a Mirax MIDI slide scanner (Zeiss, Jena, Germany) at 40x resolution (0.116 micron/pixel). Digital image analysis was performed using NearCyte software (Andrew Lesniak, University of Pittsburgh) as previously described (Krivinko et al., 2018). All analyses were done blinded to psychosis status.

### Nucleic Acid Extraction

Frozen tissue was bead-homogenized (Benchmark Scientific, Model No. D1030) in Trizol (Invitrogen; Carlsbad, CA), followed by phase separation using chloroform (20% vol:vol). RNA was then precipitated with isopropanol, washed in 75% ethanol, and resuspended in DEPC-treated water. RNA concentration and purity were assessed using Nanodrop optical density reader.

### RNA Sequencing

RNA sequencing was performed at the Next-Generation Sequencing Core at the University of Pennsylvania, Philadelphia, PA. Library preparation was performed using Illumina truSeq stranded total RNA (ribo-Zero) kits, followed by single-end, 100 bp sequencing on a HiSeq4000 sequencer. Samples from 86 AD subjects were submitted but 6 cases did not pass quality control standards for sequencing and were eliminated from further analyses. Sequencing reads were checked using FastQC and aligned to the reference genome (human genome hg38) using TopHat2 (Kim et al., 2013). RNA-seq reads that had low sequencing quality or mapping quality were filtered out. Gene expressions were then quantified using Cufflinks (Trapnell et al., 2010) and a generative statistical model of RNAseq experiments was applied to estimate FPKM (expected Fragments Per Kilobases per Million mapped fragments). A total of 22,440 genes were identified and passed initial QC. After removing genes with missing names or those not quantified in more than 50% samples in both patient groups, 15,346 genes remained (Table S1). Missing values of each remaining gene were imputed by the half of the minimum observed value across all samples. We then applied quantile normalization across samples using normalizeQuantiles function from R package Limma, and the resulting data were utilized for the downstream analyses (Mostafavi et al., 2018; Ritchie et al., 2015).

### Cell Type Fraction Estimation

We applied the est_frac function from R package MIND (Wang et al., 2020) to estimate the cell type proportions in our RNAseq data. It used the non-negative least square deconvolution approach proposed by Wang et al. (Wang et al., 2018). The signature matrix was taken from the adult single-cell RNAseq data from Darmanis et al. (Darmanis et al., 2015), which contains 666 genes and 6 cell types (astrocytes, endothelial cells, microglia, excitatory neurons, inhibitory neurons, and oligodendrocytes). The two-sample t-test was then applied to compare each cell type proportion between AD+P and AD-P samples.

### Differential Expression Analysis

Analysis of covariance (ANCOVA) was performed for each gene on the log_2_ transformed scale to compare between AD+P and AD-P, adjusted for neuropathological variables including tau area ratio, TDP-43 pathology status, HLA-DR:Iba1 ratio, microvascular lesion count and sum score. ANCOVA adjustments regarding neuropathological variables were included based on previous data which indicated an amount of predictive strength toward psychosis (Krivinko et al., 2018). Analyses also adjusted for the proportion of endothelial cells, oligodendrocytes, and excitatory neurons (which were found to be different or marginally different (p < 0.10) between AD+P and AD-P from cell type proportion estimation) (File S1 & S2). Other potential confounders (sex, APOE4, antipsychotic drug use) were evaluated for potential effects on gene expression (Figure S1). Because none of these three variables demonstrated significant main effects or interactions with psychosis impacting gene expression they were excluded from the ANCOVA models.

The covariate-adjusted fold change, p-value and FDR adjusted q-value were obtained. We further examined differentially expressed genes within each cell type by adapting CellDMC from R package EpiDISH on our RNAseq data ((Zheng et al., 2018); File S3). CellDMC was originally designed for identification of differentially methylated cell types in epigenome-wide association studies through an interaction model. We used the gene expression as the outcome and tested the interaction between cell type proportion and the psychosis status, with the same neuropathological variables adjusted.

### Correlation Analysis

Previously we reported that the mean ratio of synaptic proteins were increased in AD-P relative to AD+P (Krivinko et al., 2018). In our RNAseq data, 180 transcripts of the 190 synaptic proteins previously evaluated were present. First, we fit the same ANCOVA model (without cell type proportions adjusted) as in Krivinko et al. (Krivinko et al., 2018) for each of these 180 genes and obtained the log_2_ fold change. Then we computed the Spearman’s correlation between the previously reported protein-level fold change and the transcriptomic-level fold change for these 180 genes. We further examined whether this protein-transcriptome correlation changed after cell type proportions being adjusted in the RNAseq ANCOVA analysis.

### Gene Co-Expression Network Analysis

Gene co-expression network analysis was performed using the R package MEGENA (CRAN) (Song and Zhang, 2015). The module size was set to contain at least 15 genes. Pearson correlation was used for constructing the correlation matrix. The number of permutations for calculating FDRs for all correlation pairs and connectivity significance p-value was set to 100. After identification of modules and computing their corresponding eigengenes (MEs), we constructed a prediction model for the presence of psychosis. We first tested all MEs in individual logistic regressions, adjusting for the same neuropathological measures and three cell type proportions as covariates. The final prediction model consisted of all MEs from the individual analyses with p<0.1, and the same neuropathological variables and cell type proportions. The performance of the prediction model is characterized by area under the curve (AUC) and Nagelkerke index. We compared the nested prediction models with and without cell type proportions and/or MEs using the likelihood ratio test, as well as the DeLong’s method for comparing AUCs (DeLong et al., 1988), to assess whether including cell type proportions and MEs improve the model fitting and predictive performance.

### Functional Gene Annotation

A gene list was generated by combining all genes that were listed in individual modules which had significant association with AD+P in the multivariate analysis. This list was submitted for *Core Expression Analysis* using Ingenuity Pathway Analysis (IPA, Qiagen). Top canonical pathways with Benjamini-Hochberg p < 0.05 and number of genes ≥ 4 were identified.

## Results

### Deconvolution of Cell Type Proportions

Cell type proportion was estimated for astrocytes, endothelial cells, microglia, excitatory neurons, inhibitory neurons, and oligodendrocytes (Figure 1). Notably, there was a decreased proportion of excitatory neurons in AD+P compared to AD-P (T = 2.27; df = 75, p = 0.026). This decrease was offset by a corresponding increase in the proportion of oligodendrocytes (T = -2.17, df = 68, p = 0.033) and endothelial cells (T = -1.93, df = 78, p = 0.057). Consistent with our prior report that Iba1 volume fraction was not significantly different between AD+P and AD-P subjects (Krivinko et al., 2018), we found no change in microglia proportion (T = -0.03, df = 67, p = 0.978).

**Figure 1.**
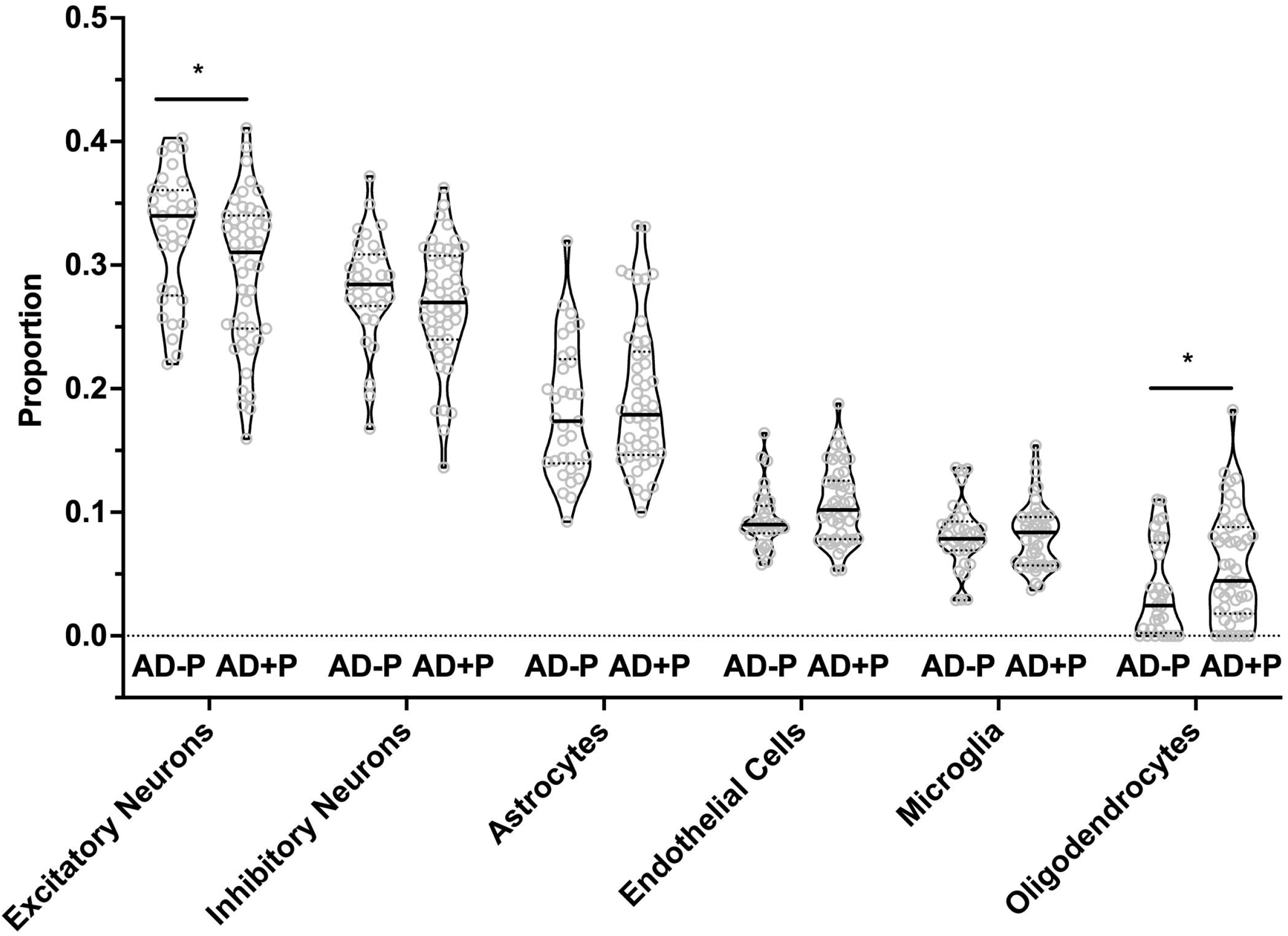
Excitatory Neurons, Oligodendrocytes, and Endothelial Cell proportions are altered in AD+P. Cell type proportion differences between AD+P and AD-P were determined using MIND signature gene matrix (Darmanis et al., 2015). Excitatory neuron proportion was significantly reduced in AD+P subjects compared to AD-P (T = 2.27 df = 75, p = 0.026). Conversely, oligodendrocyte (T = -2.17, df = 76, p = 0.033) and endothelial cell (T = -1.93, df = 78, p.= 0.057) proportions were increased in AD+P compared to AD-P. Proportions of astrocytes, microglia, and inhibitory neurons did not differ between groups (astrocyte T = -0.90, df = 73, p = 0.372 microglia T = -0.03, df = 67, p = 0.978; inhibitory neuron T = 1.40, df = 74, p = 0.166). *Indicates p < 0.05.

### Differential Expression Analysis

Our data set underwent differential expression (DE) analysis with adjustments for neuropathological covariates (Krivinko et al., 2018) and the proportions of excitatory neurons, oligodendrocytes, and endothelial cells. We identified 1077 nominally DE genes (p<0.05), across all cell types (Figure S2, Table S2A, Files S1 & S2). However, none of the 1077 genes passed an FDR threshold of q < 0.1. Additionally, we probed DE genes by cell type. Depending on cell type, we identified a range of 604 to 1287 nominally DE genes (Table S2B; File S3). Two genes passed FDR threshold: *METTL22* (q = 0.099) in Inhibitory Neurons and *DKC1* (q = 0.072) in Oligodendrocytes.

### Gene Co-Expression Network Analysis

Co-expressed genes were clustered into 288 modules (File S4). We tested each ME for strength of psychosis prediction in a multiple regression model along with neuropathologic covariates and the cell type proportions that differed between AD+P and AD-P (endothelial cells, excitatory neurons, and oligodendrocytes). We identified significant differential expression of 27 MEs between AD-P and AD+P. To avoid redundancy, parent modules that showed significant differential expression were removed, bringing our total to 23 individual modules with p < 0.05 (File S5). When these were then entered into a multivariate model only 8 modules remained significant (Table S3). Functional annotation of the 8 modules identified several significantly enriched canonical pathways (Table 2). These include regulation of protein availability via protein ubiquitination system pathways, unfolded protein response, EIF2 signaling pathways, and endoplasmic reticulum stress pathways.

**Table 2.** Functional Annotation Analysis of Differentially Expressed Gene Modules. Six significantly enriched canonical pathways were determined using IPA core expression analysis.

| Canonical Pathway | Genes Identified | p-value |
| --- | --- | --- |
| Protein Ubiquitination Pathway | ANAPC5, CRYAB, DNAJB6, DNAJC21, HSPAA1, HSPAB1, HSP90B1, HSPA4, HSPA5, HSPA4L, HSPE1, HSPH1, MED20, SUGT1, UBC, UBE2F, UBR2 | 9.21E-06 |
| Unfolded Protein Response | ATF4, DDIT3, DNAJB6, DNAJC21, HSP90B1, HSPA4, HSPA5, HSPH1, SCAP | 1.58E-04 |
| EIF2 Signaling | AGO3, ATF4, DDIT3, EIF3A, EIF4G2, EIF5B, HSPA5, RPL3, RPL8, RPL30, RPL36, RPL37, RPL26L1 | 2.57E-04 |
| Aldosterone Signaling in Epithelial Cells | CRYAB, DNAJB6, DNAJC21, HSP90AA1, HSP90AB1, HSP90B1, HSPA4, HSPA5, HSPA4L, HSPE1, HSPH1 | 2.82E-04 |
| Endoplasmic Reticulum Stress Pathway | ATF4, DDIT3, HSP90B1, HSPA5 | 5.02E-03 |
| Mitotic Roles of Polo-Like Kinase | ANAPC5, HSP90AA1, HSP90AB1, HSP90B1, RAD21 | 4.74E-02 |

### Synaptic Gene Expression

We had previously reported in a subset of our case cohort that the mean ratio of 190 synaptic proteins was greater in AD-P relative to AD+P (Figure 2A), and neuropathologic control subjects (Krivinko et al., 2018). We evaluated whether increased synaptic protein abundance in AD-P relative to AD+P subjects results from mRNA upregulation. We successfully measured the abundance of mRNAs corresponding to 180 of the 190 proteins and found that 69.4% of these mRNAs were upregulated in AD-P relative to AD+P. This pattern was not observed in non-synaptic mRNAs (Figure 2B; Chi-square test, p = 3.874E-7). However, the correlation between synaptic transcript and protein levels was modest (spearman’s rho = 0.2257, p = 0.0024, Figure 2C, File S6). Given the observed differences in cell type proportions between groups, we asked if alterations in cell type proportions might drive the differences in synaptic gene levels. When cell type proportions were introduced into quantile normalization to estimate transcript expression, there was no longer a downregulation of the synaptic genes in AD+P (Figure 2D) and transcript levels were not correlated with protein levels (spearman’s rho = 0.0302, p = 0.6868, Figure 2E, File S6).

**Figure 2.**
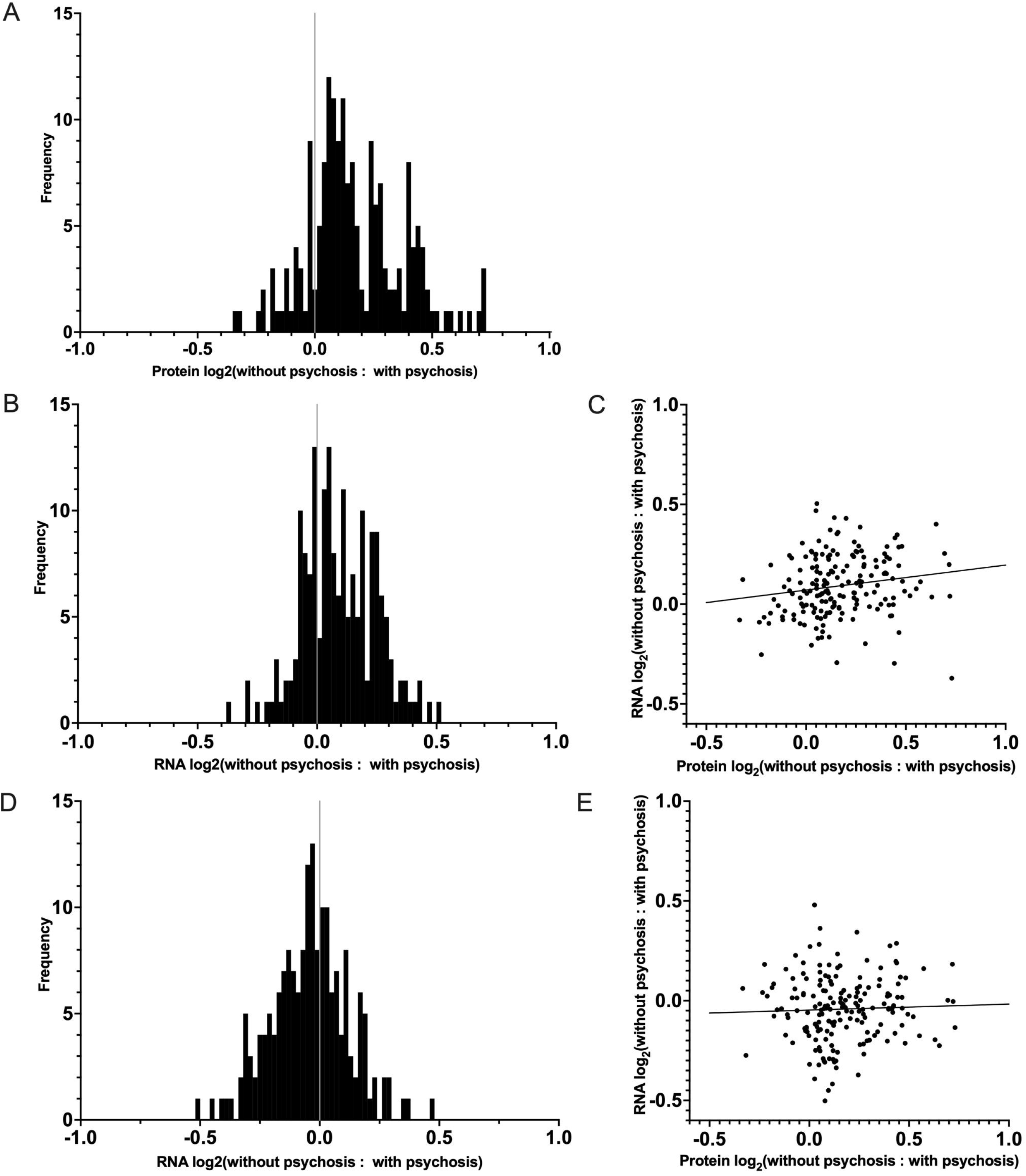
Distribution and Correlation of Synaptic Transcript and Protein levels in AD-P relative to AD+P Subjects. (A) Distributions of log_2_ ratios are shown for 180 synaptic proteins (19) for which corresponding mRNA levels were quantified in the current study. (B) Distribution of RNA expression ratios of the 180 synaptic genes identified in our DE analysis, prior to cell type proportion adjustments. The proportion of synaptic transcripts upregulated in AD-P compared to AD+P was 69.4%, compared to 49.6% of non-synaptic transcripts (Chi-square test, p = 3.874E-7) (C) Correlation between the 180 synaptic protein and transcript expression ratios (AD-P:AD+P, spearman’s rho = 0.2257, p = 0.0024). (D) Analysis of the same synaptic transcripts as in (B), accounting for the contribution of cell type proportions as covariates, eliminates their upregulation in AD-P. (E) Inclusion of cell type proportions as covariates in analysis of synaptic transcripts similarly abolishes the correlation between synaptic transcript and protein levels (spearman’s rho = 0.0302, p = 0.6868).

### Models for Prediction of Psychosis

We previously reported that a combination of neuropathologic variables (immunostaining for phospho-Tau, TDP-43, HLA-DR:Iba1 ratio, and microvascular lesion count plus a summary score of vascular pathologies) together predicted 18% of the variance in psychosis risk in AD (Krivinko et al., 2018). We first undertook to repeat that prediction in the current sample of 80 AD subjects, a subset of the subjects in our prior report. The results of these analyses are shown in Table 3. Neuropathology alone predicted 11.7% of the variance in psychosis in these subjects (Model 1). The addition of the three cell type proportions improved this prediction to 29.9% with a corresponding improvement in the area under the curve (AUC) (Model 2). Finally, entry of both the 23 significant modules and 3 cell type proportions improved prediction to 67.5% with an AUC of 0.923 (Model 3). For comparing the predictiveness among these three models, the DeLong’s AUC test shows a significant p-value for Model 2 vs Model 1 (p = 0.021), Model 3 vs Model 1 (p = 0.00014), and for Model 3 vs Model 2 (p = 0.012). On the other hand, the likelihood ratio test shows Model 2 or Model 3 fits the data significantly better than Model 1 (p = 0.006 and 0.0008, respectively), and Model 3 fits the data marginally better than Model 2 as well (p = 0.0.063).

**Table 3.**
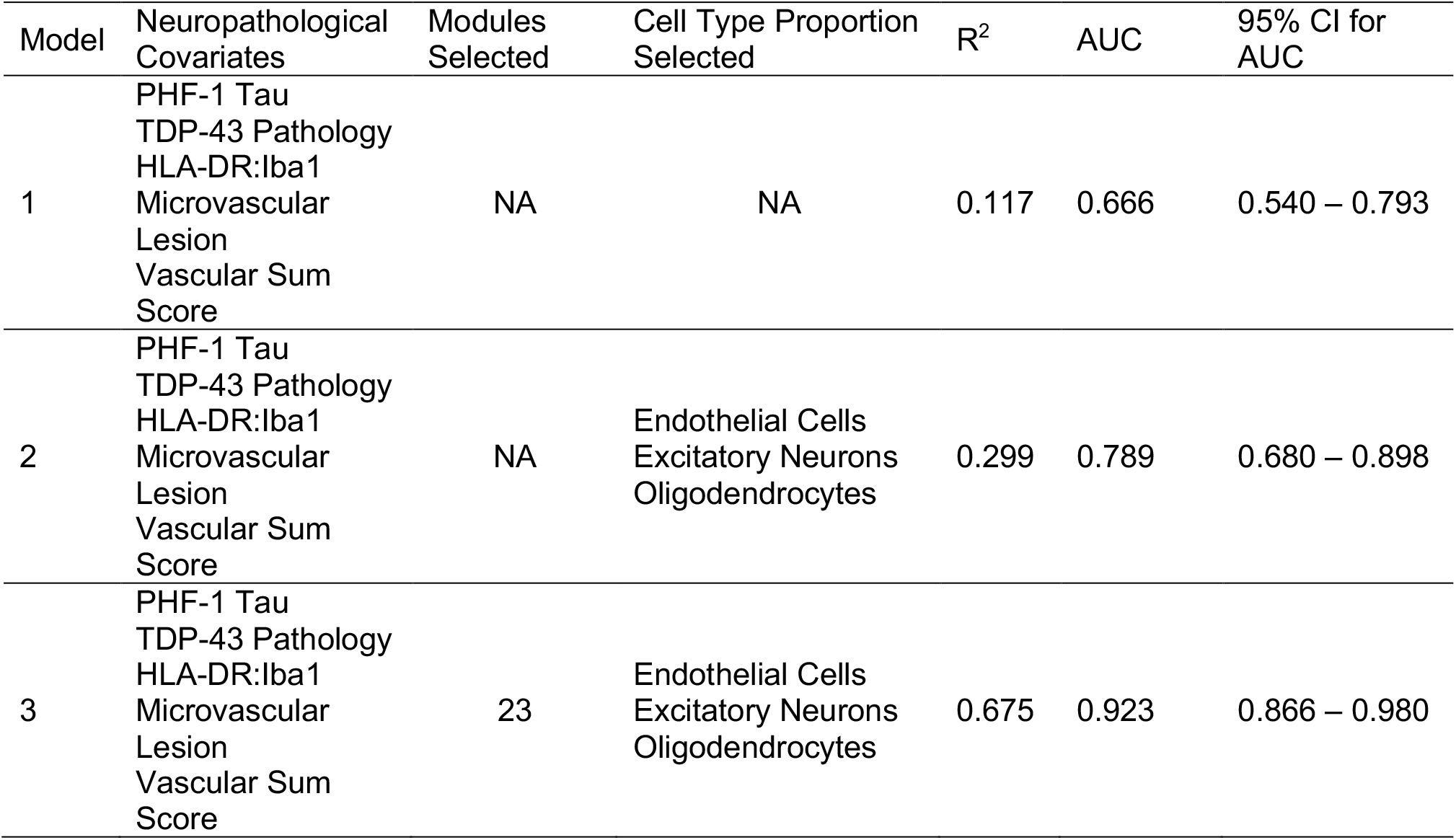
Comparison of Models for Prediction of Psychosis. Models were constructed using combinations of neuropathological covariates, cell type proportions, and co-expression modules. Model 1 tested for the prediction psychosis using only neuropathological covariates. Model 2 provided a modest improvement in predictive strength by incorporating cell type proportion estimations. Model 3, including neuropathological covariates, cell type proportions, and co-expression modules provided the greatest predictive strength.

| Model | Neuropathological Covariates | Modules Selected | Cell Type Proportion Selected | R<sup>2</sup> | AUC | 95% CI for AUC |
| --- | --- | --- | --- | --- | --- | --- |
| 1 | PHF-1 Tau<br>TDP-43 Pathology<br>HLA-DR:Iba1<br>Microvascular Lesion<br>Vascular Sum Score | NA | NA | 0.117 | 0.666 | 0.540 – 0.793 |
| 2 | PHF-1 Tau<br>TDP-43 Pathology<br>HLA-DR:Iba1<br>Microvascular Lesion<br>Vascular Sum Score | NA | Endothelial Cells<br>Excitatory Neurons<br>Oligodendrocytes | 0.299 | 0.789 | 0.680 – 0.898 |
| 3 | PHF-1 Tau<br>TDP-43 Pathology<br>HLA-DR:Iba1<br>Microvascular Lesion<br>Vascular Sum Score | 23 | Endothelial Cells<br>Excitatory Neurons<br>Oligodendrocytes | 0.675 | 0.923 | 0.866 – 0.980 |

## Discussion

We have previously shown that only a minor fraction of the risk for psychosis in AD is attributable to the severity of multiple neuropathological lesions that are present in individuals with AD. We have also previously demonstrated that a decrease in post-synaptic protein abundance was associated with psychosis in AD. Here, we set out to improve our knowledge of neurobiological contributors to psychosis in AD and the mechanisms underlying the synaptic protein decrease by evaluating the transcriptome of AD subjects with and without psychosis. Our data suggests that decreased excitatory neuron proportion contributes to underlying neurobiological perturbances associated with psychosis in AD. In addition, network analysis of the transcriptome identified a novel association with gene modules enriched for mRNAs regulating protein availability. Finally, inclusion of cell type proportions and differentially expressed module eigengenes significantly increased the predictive strength of our models beyond that of neuropathology alone, explaining 67.5% of the variance in psychosis presence.

### Excitatory neurons are vulnerable in Alzheimer’s disease with psychosis

We estimated that there was a decrease of 9% in the excitatory neuron proportion in AD+P subjects compared to AD-P, after controlling for relevant neuropathologic variants. Because these estimates are derived from measurements of levels of expressed excitatory neuron selective mRNAs, they could reflect either true cell loss or transcript downregulation. Disentangling these two possibilities will require future single-cell RNA sequencing studies of AD+P. However, several lines of evidence support the former conclusion. DLPFC neuron loss occurs in Alzheimer’s disease, including within individuals in the mild to moderate stages of disease as in the current study (Bussire et al., 2003). Similarly, levels of MAP2 protein, a marker of neuronal survival (Kirkwood et al., 2016) are lower in DLPFC of mild to moderate AD subjects relative to healthy controls (Sweet et al., 2016). Finally, in a subgroup of the current subjects, and controlling for neuropathologic burden, we reported a 32% decrease in MAP2 protein levels in AD+P subjects compared to AD-P subjects, while both AD phenotypes had significantly less MAP2 peptide levels than unaffected control subjects (Krivinko et al., 2018). Thus, it is most likely that both groups of AD subjects in the current study have excitatory neuron loss in DLPFC, and that the degree of this loss is greater in AD+P than AD-P. This interpretation would suggest that there is a modest increase in a neurodegenerative mechanism underlying psychosis in AD, beyond the degeneration due to accumulation of the measured pathologies.

### Synaptic Transcriptome

Previously, we found that AD-P subjects had an increased level of synaptic proteins compared to AD+P and control subjects (Krivinko et al., 2018). Our current analysis indicates that mRNA levels contribute only modestly to this synaptic protein compensation in AD-P relative to AD+P. Moreover, because the correlation of transcript levels with protein levels was eliminated when controlling for cell type proportions, the modest contribution of transcript abundance to synaptic protein levels seems to derive from enhanced excitatory neuron loss in AD+P, rather than from a decreased rate of transcription per se. Because synaptic proteostasis is a highly dynamic process, where the effects of transcription are modified by local translation, protein trafficking, and degradation, it is likely that these post-transcriptional mechanisms underlie synaptic protein availability in AD+P. For example, we have recently identified that genetic variation in *SUMF1*, which encodes the formylglycine generating enzyme protein, SUMF1, is associated with psychosis risk in AD in a genome-wide analysis (DeMichele-Sweet et al., 2020). SUMF1 activates multiple lysosomal sulfatases and thus genetically driven alterations in SUMF1 expression or function may influence rates of synaptic protein degradation by the lysosome to autophagosome pathway, impacting synaptic function and risk of psychosis (Amadi et al., 2020; Cosma et al., 2004; Schlotawa et al., 2020).

Additionally, our current data suggests a possible role for altered post-transcriptional mechanisms which limit synaptic protein availability in association with psychosis in AD. In our analysis of differentially expressed gene modules, protein ubiquitination pathways, unfolded protein response pathways, eukaryotic initiation factor 2 (EIF2) signaling pathways and endoplasmic reticulum stress pathways were significantly enriched (Table 2). These pathways play a critical role in regulating protein translation and degradation in neurons, including activity-dependent translation and degradation in support of synaptic plasticity (Bellato and Hajj, 2016; Costa-Mattioli et al., 2007; Lin et al., 2021; Martin and Zukin, 2006; Oliveira et al., 2021; Saito et al., 2018; Smith et al., 2020). Thus, our findings suggest that synaptic proteostasis regulation is a key player contributing towards risk of psychosis in AD.

## Conclusion

In this study, we aimed to further uncover neurobiological mechanisms contributing to psychosis in AD. Our current findings suggest that excitatory neurons are selectively vulnerable in the DLPFC of AD+P subjects relative to AD-P. Further, our findings point to the roles of post-transcriptional mechanisms underlying synaptic deficits in AD+P and identify ubiquitination, unfolded protein response, eukaryotic initiation factor 2 (EIF2) signaling, and endoplasmic reticulum stress pathways as possibly contributing to alter synaptic protein abundance in AD+P. Given that the reduction in synaptic indices in AD+P relative to AD-P subjects is common throughout the neocortex (Murray et al., 2014), the current findings, despite regional variations in gene expression and neuropathology burden, are likely to be conserved in other neocortical regions.

## Data Availability

Data will be available upon request

## Acknowledgements

This work was supported by NIH grants AG005133 (OLL, RAS, JK), AG066468 (OLL, RAS, JK) MH116046 (RAS, JK, MRD, MLM, YD, YW), K01 MH107756 (MLM).

The content is solely the responsibility of the authors and does not necessarily represent the official views of the National Institute of Mental Health, the National Institutes of Health, or the United States Government.

## Author Contributions

MRDM analyzed the RNA-sequencing data, performed functional annotation analysis, and wrote the manuscript. YW and YD performed raw data processing of RNA-sequencing data, Gene Network analysis, Cell type proportions, and differential gene expression analysis. CMW assisted with RNA sample prep and analysis. JK conducted all neuropathologic assessments and with RAS designed and oversaw all aspects of the conduct of the study. JMK and MLM provided the analyses of the synaptic proteome data. RAS and OLL contributed to the recruitment and assessment of all study subjects while alive. All authors reviewed and approved the final manuscript.

## Supplemental Methods

## Appendix 1. Subjects

We studied a total of 80 AD subjects (Table 1) obtained through the brain bank of the Alzheimer Disease Research Center (ADRC) at the University of Pittsburgh, using protocols approved by the University of Pittsburgh Institutional Review Board and Committee for Oversight of Research and Clinical Training Involving Decedents. All cases coming to autopsy between 1993 and 2014 with a primary neuropathologic diagnosis of Alzheimer’s disease and a Braak stage between 3-5 were included in the study. End-stage cases, as defined by a Braak stage of 6, were excluded, as clinically the greatest increase in onset of psychosis in AD occurs between early and middle disease stages (Lopez et al., 2003; Ropacki and Jeste, 2005).

## Appendix 2. Sample Collection and Neuropathological Assessment

For ADRC subjects, postmortem interval (PMI) was recorded at the time of brain removal. At autopsy, the brain was removed intact, examined grossly, and divided in the midsagittal plane. Gray matter samples from the right superior frontal gyrus of the DLPFC were dissected and frozen at -80 °C. The left hemibrain was immersion fixed in 10% buffered formalin for at least one week, sectioned into 1.0 cm coronal slabs, and sampled according to Consortium to Establish a Registry for Alzheimer’s Disease (CERAD) protocol for neuropathological diagnosis of AD (Mirra et al., 1991) or, since 2012, following National Institute of Aging – Alzheimer’s Association (NIA-AA) guidelines (Montine et al., 2012). AD pathology was evaluated using the modified Bielschowsky silver stain (Yamamoto and Hirano, 1986) and immunohistochemical staining for tau and amyloid β. Neuritic plaque density was assessed according to CERAD criteria (Mirra et al., 1991); distribution of tau pathology was classified according to Braak stages (Braak et al., 2006). Neuropathologic diagnoses of Alzheimer disease were made according to NIA-Reagan criteria (Hyman and Trojanowski, 1997), with all cases meeting criteria for intermediate to high probability that their dementia was due to AD. Lewy body pathology was initially assessed in amygdala, brainstem and olfactory bulb, and if positive, further evaluated in limbic and neocortical sections following consensus guidelines (McKeith et al., 2017; Montine et al., 2012). Immunohistochemical staining for phospho-TDP-43 was performed on sections of amygdala, hippocampus, mesial temporal cortex and middle frontal gyrus as previously described (Vatsavayi et al., 2014). Sections were evaluated for the absence or presence of TDP-43 positive neuronal cytoplasmic inclusions, neuronal intranuclear inclusions and dystrophic neurites. Since our previous analyses did not reveal any associations of AD+P with disease stages of Lewy body or TDP-43 pathology (Krivinko et al., 2018), we continued to stratify these two proteinopathies as positive or negative, whereby all cases with any level of pathology were classified as positive and cases with complete absence of either one of these proteinopathies as negative.

Assessment of vascular pathology included atherosclerosis of the circle of Willis, arteriolosclerosis in frontal white matter and cerebral amyloid angiopathy in DLPFC. Each was rated as none (0), mild (1), moderate (2) or severe (3), and a sum score was generated by adding the three individual scores. Microvascular lesions (MVL) were defined as remote microinfarcts/microhemorrhages not seen on gross examination and less than 1.0 cm in size. MVLs were enumerated in standardized sections (Montine et al., 2012) of middle frontal gyrus (DLPFC), superior and middle temporal gyrus, inferior parietal lobule, occipital cortex (BA 17/18), basal ganglia at level of anterior commissure, and thalamus at the level of the subthalamic nucleus to create MVL counts.

## Appendix 3. Quantitative Immunohistochemistry and digital image analysis

Neuropathological disease burden in the DLPFC was previously assessed in all 80 cases using quantitative immunohistochemistry (Krivinko et al., 2018). In short, serial 5 μm thick formalin-fixed, paraffin-embedded tissue sections were immunostained on an automated stainer (Discovery Ultra, Ventana, Tucson, AZ) using the following primary antibodies: PHF-1 (1:1000, kindly provided by Peter Davies), oligomeric tau T22 (1:500, EMD Millipore, Billerica, MA), beta-amyloid NAB228 (1:4000, Cell Signaling Technology, Danvers, MA, after 40 min pretreatment with 90% formic acid), and microglial markers Iba1 (Ionized calcium binding adaptor molecule 1) (1:500, Wako, Richmond, VA) and HLA-DR (Human Leukocyte Antigen – DR isotype) (1:100, Dako, Agilent Technologies, Santa Clara, CA). Except for beta-amyloid, slides for all other stains were pretreated with Discovery CC1 solution, a Tris based buffer with a slightly basic pH (Ventana Medical Systems, Tucson, AZ). All slides were developed using a multimeric HRP/DAB detection system (Ventana Medical Systems, Tucson, AZ). No counterstaining was performed to ease signal quantification.

Whole slide digital images of the immunostained sections were created using a Mirax MIDI slide scanner (Zeiss, Jena, Germany) at 40x resolution (0.116 micron/pixel). Digital image analysis was performed using NearCyte software (Andrew Lesniak, University of Pittsburgh). For each section, 4 rectangular regions of interest (ROI) of 4mm^2^ were created. These ROIs were defined to span the entire cortical thickness and were preferentially placed midway along the gyral axis to avoid tangentially cut cortical regions. Minor manual adjustments were made to adapt to curvatures and irregularities in the cortical ribbon. Once placed for the first analyzed stain (PHF-1), the same ROIs were re-used for all subsequent stains. If tissue folds or other artifacts prevented placement in the same location, the ROI was moved to an acceptable site as close as possible to the original location. For quantitative image analysis, thresholds for signal positivity were optimized manually for each stain and then maintained constant throughout the analysis of all slides. Signals from all four ROIs were integrated into two outcome variables: area ratio (= positive area/entire field area) and mean signal intensity. For HLA-DR and Iba1 stains, an additional variable, the HLA-DR/Iba1 ratio was derived to normalize microglial activation (HLA-DR) to microglial density (Iba1). All analyses were done blinded to psychosis status.

## Supplemental Legends for Tables and Figures

**Supplemental Table 1.**
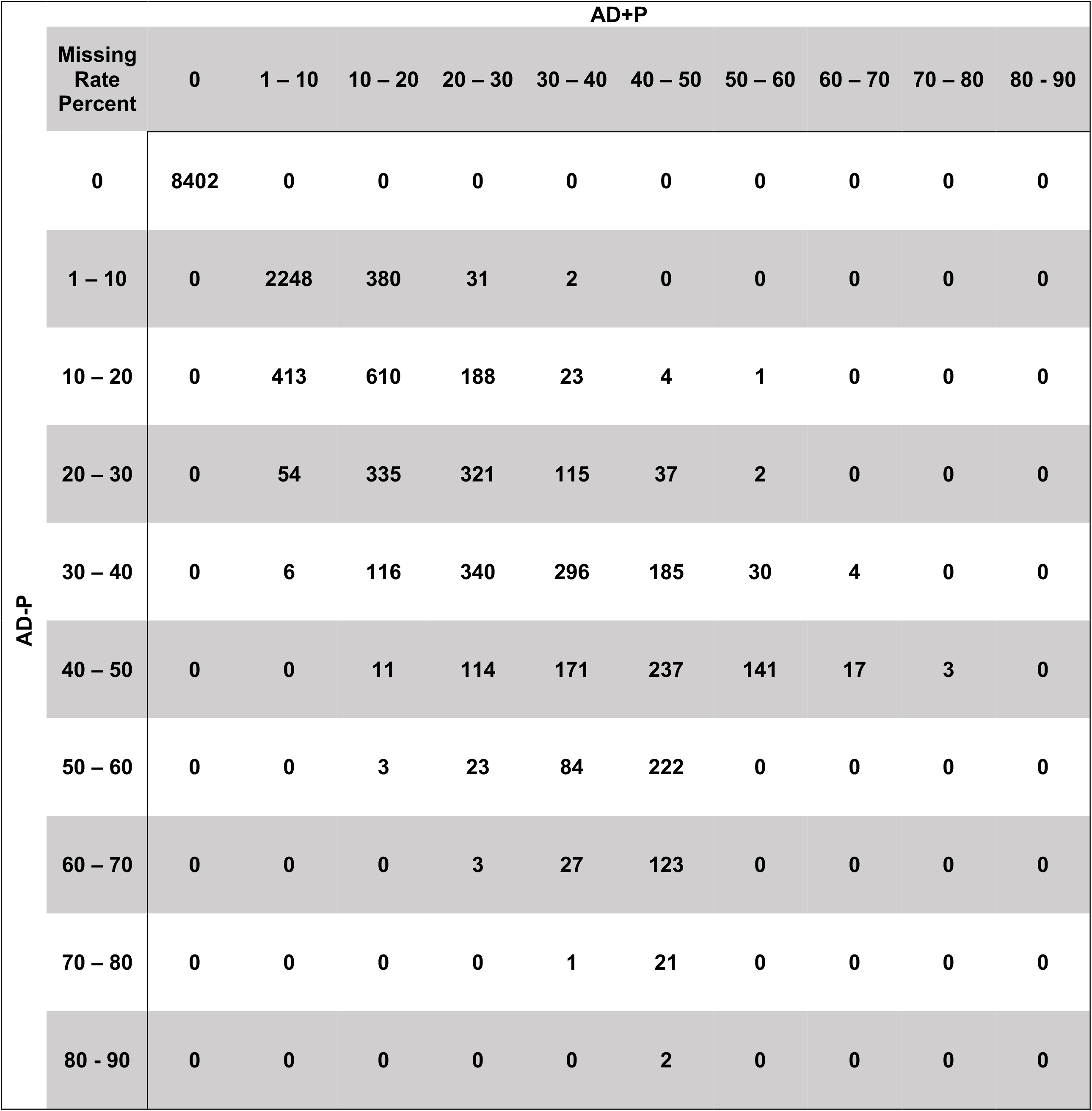
Missing Rate of Genes Retained for Analysis. RNA Sequencing identified 22.4040 genes, which were then filtered based on missing rate criteria. Among the 15,346 that remained and included for analysis, 198 genes had greater than 50% missing rate in AD+P, but less than 50% in AD-P; 509 genes had greater than 50% missing rate in AD-P, but less than 50% in AD+P. Within the set of genes included for analysis, 14,639 genes have missing rates of less than 50% for AD-P and AD+P groups.

| AD-P | AD+P |  |  |  |  |  |  |  |  |  |  |
| --- | --- | --- | --- | --- | --- | --- | --- | --- | --- | --- | --- |
|  | Missing<br>Rate<br>Percent | 0 | 1 – 10 | 10 – 20 | 20 – 30 | 30 – 40 | 40 – 50 | 50 – 60 | 60 – 70 | 70 – 80 | 80 - 90 |
|  | 0 | 8402 | 0 | 0 | 0 | 0 | 0 | 0 | 0 | 0 | 0 |
|  | 1 – 10 | 0 | 2248 | 380 | 31 | 2 | 0 | 0 | 0 | 0 | 0 |
|  | 10 – 20 | 0 | 413 | 610 | 188 | 23 | 4 | 1 | 0 | 0 | 0 |
|  | 20 – 30 | 0 | 54 | 335 | 321 | 115 | 37 | 2 | 0 | 0 | 0 |
|  | 30 – 40 | 0 | 6 | 116 | 340 | 296 | 185 | 30 | 4 | 0 | 0 |
|  | 40 – 50 | 0 | 0 | 11 | 114 | 171 | 237 | 141 | 17 | 3 | 0 |
|  | 50 – 60 | 0 | 0 | 3 | 23 | 84 | 222 | 0 | 0 | 0 | 0 |
|  | 60 – 70 | 0 | 0 | 0 | 3 | 27 | 123 | 0 | 0 | 0 | 0 |
|  | 70 – 80 | 0 | 0 | 0 | 0 | 1 | 21 | 0 | 0 | 0 | 0 |
|  | 80 - 90 | 0 | 0 | 0 | 0 | 0 | 2 | 0 | 0 | 0 | 0 |

**Supplemental Table 2A, B.** Tabulation of Differential Gene Expression Results. Table 1A displays the number of nominally significant genes binned by p-value. No genes passed the FDR threshold, q < 0.1. Table 1B shows the differentially expressed genes in each cell type binned by p-value. A large number of genes reached nominal significance in each cell type, however, only two genes passed the FDR threshold, q < 0.1: *METTL22* (inhibitory neurons, q = 0.099) and *DKC1* (oligodendrocytes, q = 0.072).

Supplemental Table 2A and 2B. Tabulation of Differential Gene Expression Results.
| A. Differentially Expressed Genes Analyzed after Quantile Normalization |  |  |  |  |  |
| --- | --- | --- | --- | --- | --- |
| p-value | $< 1^{-5}$ | $< 1^{-4}$ | $< 1^{-3}$ | $< 1^{-2}$ | $< 5^{-2}$ |
| DE Genes | 0 | 1 | 16 | 228 | 1077 |

| B. Distribution of Differentially Expressed Genes by Cell Type and p-value |  |  |  |  |  |
| --- | --- | --- | --- | --- | --- |
| p-value | $< 1^{-5}$ | $< 1^{-4}$ | $< 1^{-3}$ | $< 1^{-2}$ | $< 5^{-2}$ |
| Astrocyte | 0 | 0 | 27 | 252 | 1117 |
| Endothelial Cell | 1 | 1 | 24 | 262 | 1287 |
| Microglia | 0 | 0 | 13 | 145 | 728 |
| Excitatory Neuron | 0 | 2 | 10 | 127 | 604 |
| Inhibitory Neuron | 1 | 2 | 9 | 143 | 635 |
| Oligodendrocyte | 1 | 1 | 12 | 145 | 706 |

**Supplemental Table 3.**
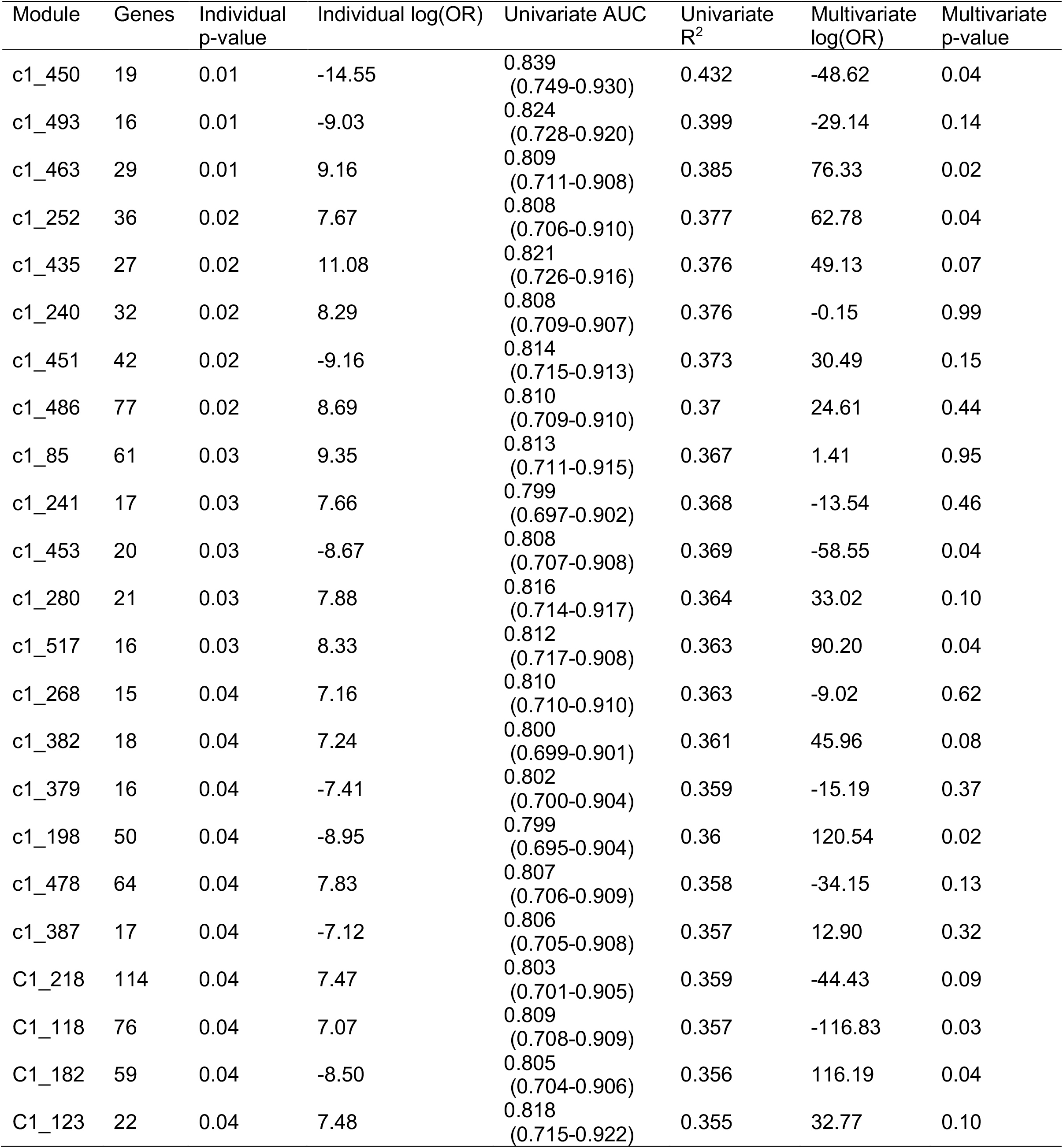
MEGENA Module Differential Expression Analysis. Quantile Normalized RNA-sequencing data was clustered and organized into 288 modules. Modules underwent univariate and multivariate logistical regression evaluation to test for predictive strength of psychosis.

Supplemental Figure 1. The Effects of Covariates on Gene Expression Are Not Significant. We tested the covariates (sex, left column; APOE E4, middle column; antipsychotic use, right column) in the following model: RNA expression ∼ confounder + psychosis + confounder-by-psychosis interaction. Top row shows the p values for the main effect of each confounder. Bottom row shows the p value for their interaction effects.

Supplemental Figure 2. Distribution of DE Genes in AD-P relative to AD+P Subjects. DE gene results underwent quantile normalization utilizing a regression model which incorporated neuropathological covariates and proportions of endothelial cells, excitatory neurons, and oligodendrocytes. The total number of nominally significant (p < 0.05) DE genes remaining after quantile analysis was limited to 1077, none of which had q < 0.1.

Supplemental File 1. Differentially expressed transcripts without cell type proportion adjustment. Transcripts are organized with respective Log2 Fold Change (Log2FC), nominal significance (p-value), the difference of RNA transcript levels (Log2) between AD+P and AD-P, and false discovery rate (q-value). Log2FC is a function of AD-P relative to AD+P. Quantile normalization was applied across samples using normalizeQuantiles function from R package Limma.

Supplemental File 2. Differentially expressed transcripts with cell type proportion adjustment. Transcripts are organized with respective Log2 Fold Change (Log2FC), nominal significance (p-value), the difference of RNA transcript levels (Log2) between AD+P and AD-P, and false discovery rate (q-value). Log2FC is a function of AD-P relative to AD+P. Quantile normalization was applied across samples using normalizeQuantiles function from R package Limma.

Supplemental File 3. Differentially expressed transcripts between cell types. Application of est_frac from R package MIND was used to estimate cell type proportion, in which transcripts found within each cell type was evaluated for differential expression.

Supplemental File 4. Supplemental File 4. MEGENA Gene Modules. Gene co-expression network analysis was performed using R package MEGENA. The minimum module size was set to 15 genes. Co-expressed genes were clustered into 288 modules.

Supplemental File 5. MEGENA Module Strength of Association with Psychosis. Module eigengenes (MEs) were first tested independently for strength of psychosis prediction (File S5A). A multiple regression model then evaluated the association between MEs and neuropathologic covariates (File S5B). Finally, these analyses were repeated by adding cell type proportions into the logistic regression as covariates (File S5C).

Supplemental File 6. Correlation between synaptic proteins and transcripts. Synaptic genes that overlapped from this study and Kivinko et al. (2018). The RNAseq data set was first adjusted using the same ANCOVA as in Krivinko et al. The protein-transcriptome correlation was then evaluated after cell type proportions were adjusted in the RNAseq ANCOVA analysis.

