## Supplementary figures and images for "Psychosis in Alzheimer’s Disease is Associated with Excitatory Neuron Vulnerability and Post-Transcriptional Mechanisms Altering Synaptic Protein Levels"

### Supplemental Figure 1

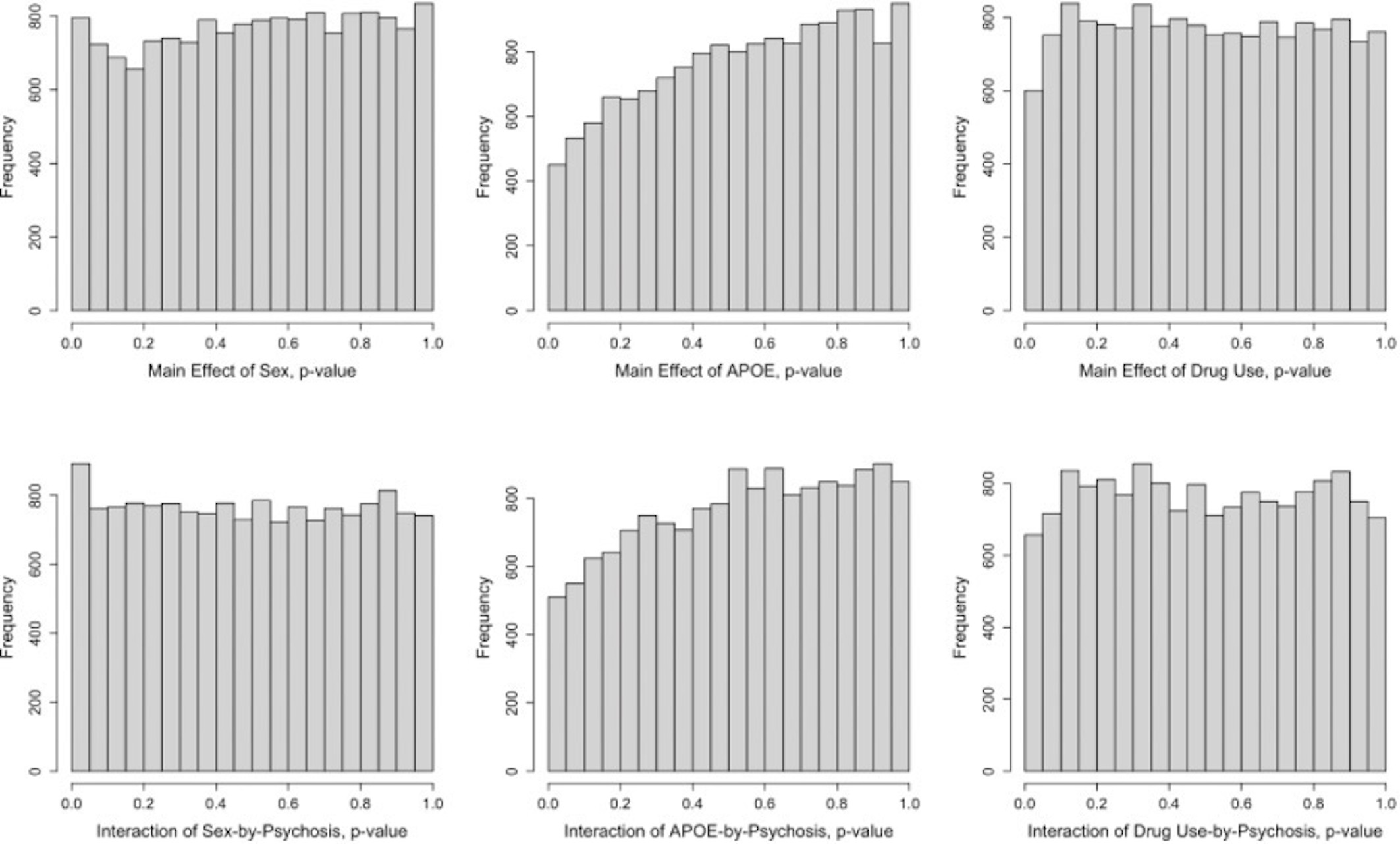

### Supplemental Figure 2

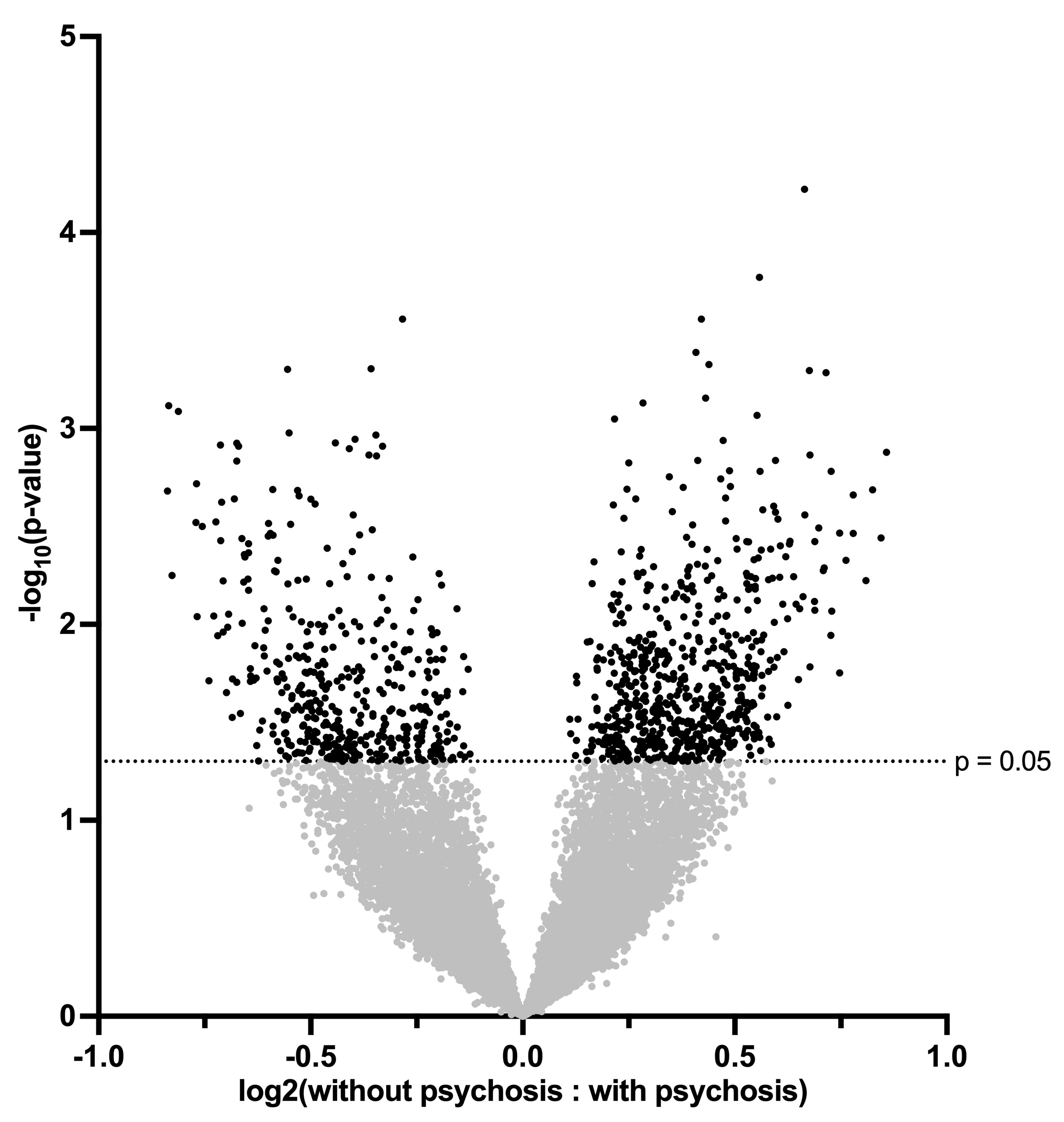
